# Sectoral Differences in Pediatric Antibiotic Prescribing for Acute Otitis Media

**DOI:** 10.64898/2026.06.25.26356579

**Authors:** Janina Hansas, Péter Csonka, Manela Karunadasa-Visama, Pekka Vartiainen, Anna-Leena Vuorinen

**Author notes:** **Address correspondence to:** Janina Hansas.

## Abstract

**Importance:** Acute otitis media is the most common infection in children and a major reason for antibiotic prescriptions, up to one third of which may be unnecessary. Sector of care may influence AOM management through differences in access to care, specialist involvement, parental expectations and financial foundation.

**Objective:** The objective is to examine differences in antibiotic prescribing practices between healthcare sectors.

**Design:** This is a nationwide register-based study comparing data from different healthcare sectors.

**Setting:** Finnish primary and secondary healthcare, covering both public- and private-sector visits. Prescriptions and sociodemographic information were linked from nationwide registers.

**Participants:** We included children under 18 years old who received a diagnosis of acute otitis media, defined by ICD-10 codes H65–H67, between January 1, 2017 and December 31, 2022.

**Exposures:** The exposure is the sector of care (public sector vs. private sector).

**Main Outcomes and Measures:** Primary outcomes were antibiotic prescribing, guideline adherence of the prescribed antibiotics, and rates of management failure. Secondary outcomes included antibiotic selection and guideline-adherent eligibility for tympanostomy tube placement. Associations were estimated using adjusted odds ratios (aORs) with 95% confidence intervals (CIs).

**Results:** The study included 295 064 children with 596 634 acute otitis media index visits, of which 77.6% resulted in an antibiotic prescription. Private-sector visits were associated with higher odds of antibiotic being prescribed (adjusted odds ratio [aOR]: 1.45; 95% CI: 1.41–1.49).

Overall, 87.3% of antibiotic prescriptions were guideline adherent, but private-sector care was associated with lower odds of guideline-adherent prescribing (aOR: 0.64; 95% CI: 0.60–0.69). Compared with amoxicillin, the private sector showed higher odds of prescribing amoxicillin–clavulanic acid (32.8% vs. 8.3%; aOR: 3.00; 95% CI: 2.91–3.10).

Management failure occurred in 7.0% of episodes and was more common in the private sector (aOR:1.52; 95% CI: 1.48–1.56). Only 48.7% of all tympanostomy tube insertions met the eligibility criteria.

**Conclusions and Relevance:** In this study overall adherence to guideline-recommended antibiotic treatment for AOM was high in Finland. Nevertheless, observed clinically meaningful sectoral differences in antibiotic selection, treatment failure, and tympanostomy eligibility adherence indicate a need for targeted antimicrobial stewardship and quality-improvement efforts, especially in the private sector.

**Key Points:** *Question:* Does the healthcare sector of care influence the management of acute otitis media?

*Findings:* In this nationwide, episode-based register study of 295 064 Finnish children with acute otitis media, overall adherence to guideline-recommended antibiotic therapy was high. However, compared with the public sector, private-sector care was associated with higher odds of antibiotics being prescribed, lower guideline adherence and higher rates of management failure.

*Meaning:* Sectoral differences in care suggest opportunities to improve guideline adherence, antimicrobial stewardship, and quality of care particularly in private sector.

## INTRODUCTION

Acute otitis media (AOM) is the most common infection among children in outpatient care and a major driver of antibiotic use, yet up to one third of antibiotic prescriptions may be unnecessary given its high self-resolution rates.^1–4^ Antibiotic overuse, particularly the use of broad-spectrum agents, contributes to antimicrobial resistance, which is linked to over a million deaths annually and increases the risk of adverse effects, including *Clostridium difficile* infection and long-term disruption of the childhood microbiome.^2–6^

Despite established guidelines recommending watchful waiting in most cases and reserving antibiotics for unequivocal diagnoses or high-risk situations,^7^ adherence to recommended practices varies significantly across and within countries. Deviations commonly involve inappropriate antibiotic selection or suboptimal dosing.^8–10^

Finland’s, primary care is delivered through both publicly and privately funded services (eMethods in **Supplement 1**), which may influence clinical decision-making, including prescribing and management practices. Despite universal access to publicly funded care, a substantial proportion of children (40%-60%) are covered by private health insurance.^11^ Sector of care may influence AOM management through differences in in access to care, specialist involvement, parental expectations and financial foundation.^11^ Examining sectoral variation is therefore important to distinguish differences that reflect patient case mix from those related to healthcare system factors.

Private healthcare providers may more likely prescribe antibiotics and broad-spectrum agents for respiratory infections than public providers, and they also tend to prescribe more expensive medications.^12–16^ However, while sectoral differences are recognized, no prior studies have examined sector-specific AOM management at a population level using a consistent, episode-based approach.

Using nationwide register data from 2017 to 2022, we investigated whether the management of AOM—including prescribing antibiotics, guideline adherence, treatment failure, and eligibility for tympanostomy tube insertion—differed between the public and private healthcare sectors in Finland.

## METHODS

### Design and data sources

This is a nationwide register-based study linking individual-level data from primary and secondary healthcare registers, ^17^ national Prescription Centre data,^18^ and population registers.^19–21^ Detailed descriptions of data sources are provided in eMethods **Supplement 1**.

The registers include both public and private healthcare providers; private sector data have been available since 2015, with near-complete coverage during the later study years.^22^ Data permits were received from the Finnish Social and Health Data Permit Authority (THL/2698/14.02.00/2023 as amended) and Statistics Finland (TK/1424/07.03.00/2024 as amended).

### Participants

We included all children under 18 years of age who had at least one healthcare contact recorded in the Finnish primary or secondary care registers between January 1, 2017, and December 31, 2022, and received a diagnosis of AOM (diagnosis codes H65–H67 ; eMethods in **Supplement 1**) as defined by the International Classification of Diseases, 10th Revision (ICD-10). The tympanostomy sub-cohort comprised all main-cohort children who underwent tympanostomy tube insertion (Nordic Classification of Surgical Procedures, NCSP, procedure codes SPAT1019, DCA20) between January 1, 2020, and December 31, 2022. We used this study period to ensure better coverage of private sector visits.

### Data processing

We identified AOM visits using ICD-10 codes H65–H67 and linked them to sociodemographic and Prescription Centre data as described in the eMethods; eFigure 1 (**Supplement 1**). We considered antibiotic prescriptions (ATC code J01*) issued within 2 days of a visit AOM-related (**Figure 1B**); we also evaluated 0- and 7-day windows in sensitivity analyses.

**Figure 1.**
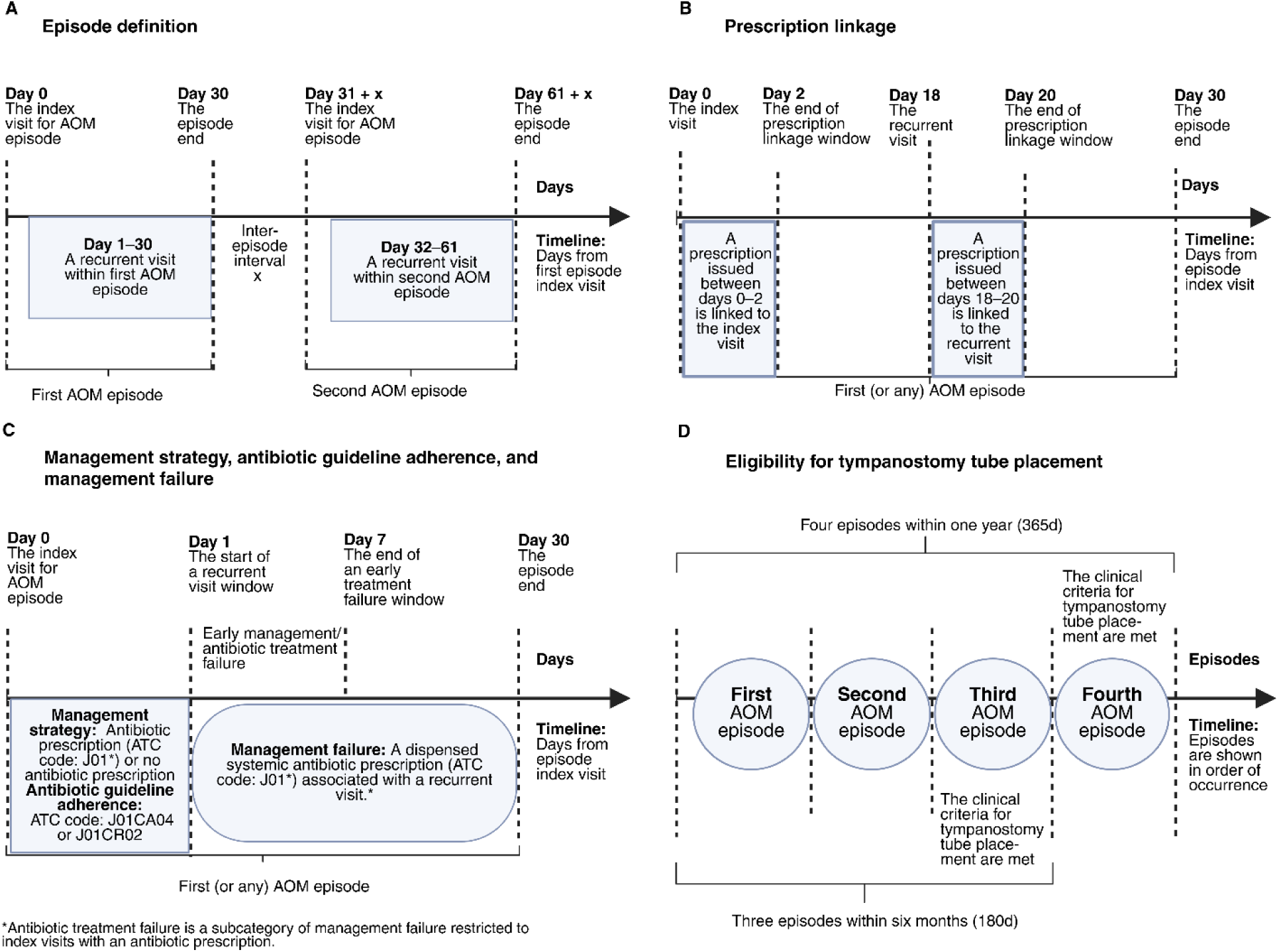
The temporal definitions and clinical criteria used to identify acute otitis media (AOM) episodes, recurrent visits, prescription linkage, treatment failure, guideline adherence, and tympanostomy tube placement eligibility. The time windows are based on national guidelines^7^ and adapted for register-based analysis. **(A)** Definitions of AOM episodes and recurrent visits. **(B)** The timeline for prescription linkage. **(C)** Definitions of management failure and antibiotic guideline adherence. We defined early management failure as occurring within 7 days of the index visit. **(D)** Time-based eligibility for tympanostomy tube insertion. To account for potential delays in the timing of the procedure, we inserted an additional 30-day window into both the years, and six months’ time demands in calculations. (Created in https://BioRender.com).

We grouped visits into episodes, defined by an index visit (the first visit or any visit ≥30 days after a preceding episode), with recurrent visits within 30 days assigned to the same episode (**Figure 1A**); 14- and 45-day windows were evaluated in sensitivity analyses. We identified tympanostomy tube placements using NCSP codes DCA20 and SPAT1019 and linked them to sociodemographic data using the same approach.

### Outcomes

#### Management strategy

We defined management strategy as *no antibiotics* vs. *antibiotics* (J01*) based on the ATC code recorded at the index visit.

#### Management failure

We defined *management failure* as a recurrent AOM visit with a systemic antibiotic prescribed and dispensed within 30 days of the index visit, regardless of antibiotic treatment at the index visit and *early management failure* identically but within 7 days (**Figure 1C**). We assigned failures to the index-visit sector; in a sensitivity analysis, we assigned them to the sector prescribing antibiotics at failure.

#### Antibiotic guideline adherence

We classified prescribing antibiotics as *guideline adherent* (prescribing amoxicillin [J01CA04] or amoxicillin-clavulanic acid [J01CR02]) vs. *non-first-line antibiotics* (reference category: other J01*), based on the ATC code recorded at the index visit.

#### Antibiotic treatment strategy

We categorized prescribed antibiotics at the index visit into a three-level variable: *amoxicillin-clavulanic, amoxicillin* (reference category), or *non-first-line antibiotics*.

We further categorized *Non-first-line antibiotics* as *second-line antibiotics* (trimethoprim-sulfa [J01EE*], macrolides [J01FA*], 2^nd^–3^rd^ generation cephalosporines [J01DC*, J01DD*]), *potentially harmful antibiotics* (fluoroquinolones [J01M*], aminoglycosides [J01GB*], tetracyclines [J01AA*]), and *suboptimal antibiotics* (all the other antibiotics in the J01 class; eMethods **supplement 1**). ^7^

#### Antibiotic treatment failure

We defined *antibiotic treatment failure* as a management failure among episodes treated with a systemic antibiotic at the index visit and *early antibiotic treatment failure* similarly but within 7 days. (**Figure 1C**).

#### Eligibility for tympanostomy

We considered procedure guideline adherent if the child had ≥3 episodes within a six month period or ≥4 within one year (**Figure 1D**), consistent with national guidelines.^7^ To account for potential delays in the procedure’s timing we extended the time window used in the calculation by 30 days.^23^ This outcome was defined as *criteria met* vs. *criteria not met*.

### Covariates

We calculated age at the visit from the date of birth and categorized as <2, 2–4.9, 5–11.9, or 12–17.9 years. We coded sex as girl or boy. Parental occupational status was based on the higher-status parent and categorized as *upper-level non-manual*, *other forms of employment* (including retirees, *n* = 5162), *unemployed, student*, or *unknown*. We categorized native language as *Finnish, Swedish*, or *other* (proxy for non-domestic-language background).

We derived chronic conditions from ICD-10 codes recorded on or before the visit, including allergic rhinitis (J30), atopic eczema (L20), immunological deficiencies (D80–D89), Down syndrome (Q90), and asthma (J45–J46).^24–26^

We defined recurrent otitis media as ≥3 episodes within a six-month period or ≥4 within one year and treated as a background factor for one year after the last qualifying episode. We categorized healthcare sector as *public* or *private* by provider type.

We classified doctor’s specialty, per the Finnish Supervisory Agency^20^, as *otorhinolaryngologist, general practicer, pediatrician, other specialty,* or *no specialty*.

We included diagnosis category as a covariate, grouping diagnoses into *H66* and *H65/H67*. Finally, season was assigned from the month of the visit.^28^

More detailed information in eMethods (**Supplement 1**).

### Statistical Analysis

We analyzed the association between healthcare sector and outcomes using generalized linear mixed-effects models (GLMMs) to account for repeated observations within individuals.

We analyzed binary outcomes with logistic mixed-effects models and the three-level antibiotic treatment strategy outcome with a Bayesian multilevel model (BMM). GLMMs accounted for within-patient clustering, while BMM parameters were estimated using Markov chain Monte Carlo, and summarized by median odds with 95% credible intervals (eMethods in **Supplement 1**).

Crude models included healthcare sector as the main exposure and a patient-level random intercept. Reference categories were: *public sector* (exposure); *no prescription* (management strategy); *other* (guideline adherence); *amoxicillin* (treatment strategy); *no recurrence* (management-failure outcome); and *indication not met* (tympanostomy eligibility).

Adjusted models included the following covariates: age, sex, physician specialty, occupational status of a parent, native language, diagnosis category, season, and medical history (selected chronic conditions and recurrent AOM; the latter excluded from the tympanostomy eligibility assessment). Reference categories were: *<2 years* (age), *boy* (sex), *no specialty* (physician specialty), *student* (occupational status), *Finnish* or *Swedish* (native language), combined *H65*/*H67* (diagnosis category), *summer* (season), and *no relevant medical history*.

We performed sensitivity analyses for management strategy and management failure using alternative episode windows, prescription-linkage windows, and an alternative definition assigning management failure to the sector issuing the antibiotic prescription. For tympanostomy eligibility, we additionally evaluated a 14-day episode window (eMethods in **Supplement 1**).

We performed all the analyses in R 4.5.1. The GLMMs were fitted using the glmmTMB package, and BMMs were implemented using the brms package.^28,29^ Codes are available in: https://github.com/HDS-DigitalEpi/Acute-Otitis-Media/tree/main)

## Results

### Cohort characteristics

The final cohort included 295 064 patients, 737 322 visits and 520 567 antibiotic prescriptions (**Figure 2**) of which 464 368 (89.2%) were dispensed. Both sectors were involved, via a visit or prescription, in 10 779 (1.81%) index visits, with antibiotics prescribed, by both in 4766. In total, 7207 children underwent 8304 tympanostomy tube placement procedures. The cohort characteristics are presented in **Table 1**. Compared to public-sector patients, patients in private sector were younger, had more often a history of recurrent AOM and they had higher occupational status.

**Figure 2.**
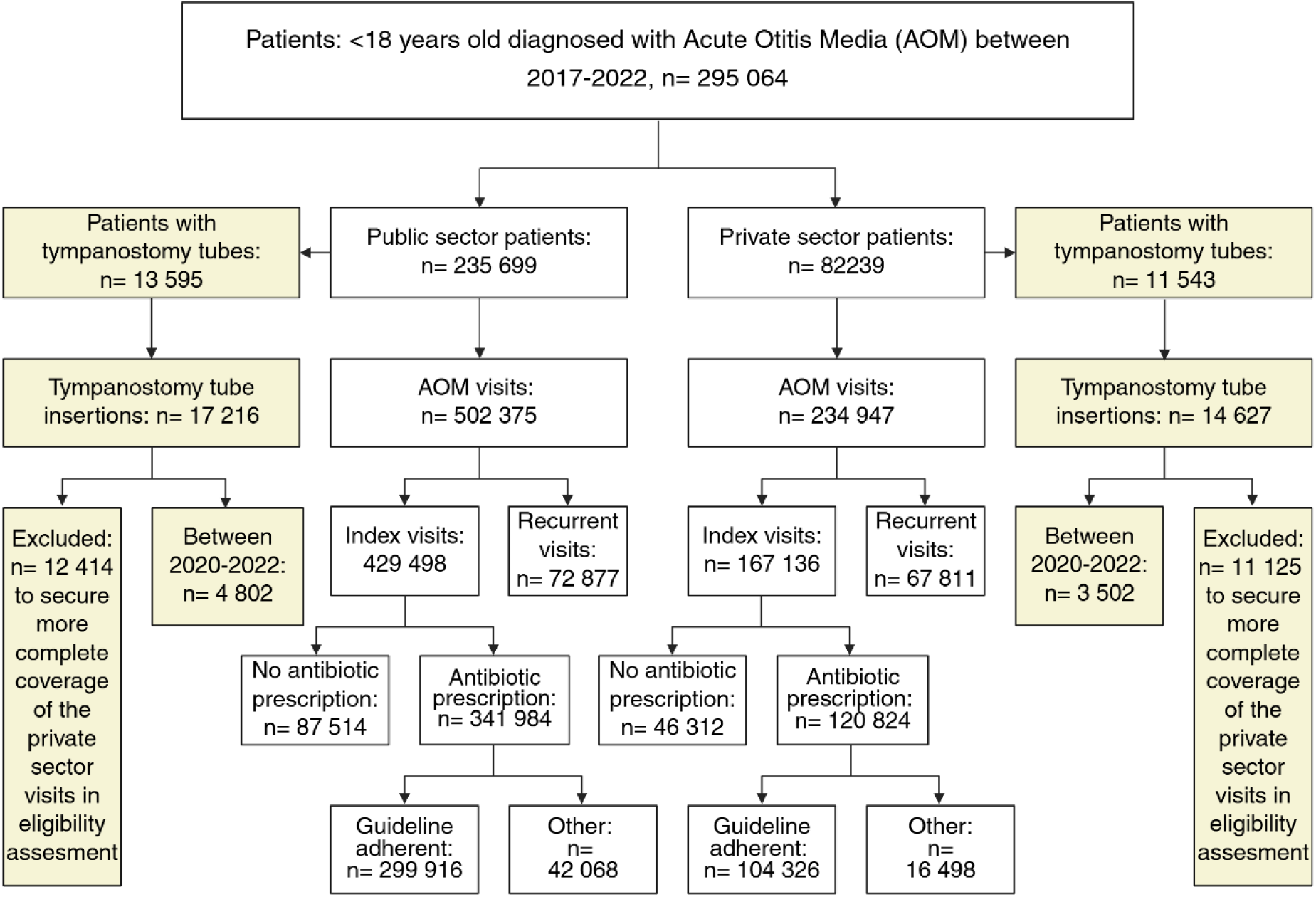
A flowchart of the retrospective cohort used to assess clinical outcomes (Created in https://BioRender.com).

**Table 1.** The background characteristics of the cohort by the first visit of an episode.

|  | Both Sectors | Public Sector No. (%) | Private Sector No. (%) |
| --- | --- | --- | --- |
| **Episodes** | **596 634** | **429 498** | **167 136** |
| Patients | 295 064 | 235 699 | 82 239 |
| Visits | 737 322 | 502 375 | 234 947 |
| Episodes per patient (mean) | 2.0 | 1.8 | 2.0 |
| Visits per patient (mean) | 2.5 | 2.1 | 2.9 |
| Age median (iqr) | 3.3 (1.7-6.1) | 3.7 (1.8-6.8) | 2.7 (1.6-4.7) |
| **Age group** |  |  |  |
| 0-2 years old | 167 321 (28.0) | 115 178 (26.8) | 52 143 (31.2) |
| 2-5 years old | 217 637 (36.5) | 145 777 (33.9) | 71 860 (43.0) |
| 5-12 years old | 168 069 (28.2) | 131 452 (30.6) | 36 617 (21.9) |
| 12-18 years old | 43 607 (7.3) | 37 091 (8.6) | 6 516 (3.9) |
| **Sex** |  |  |  |
| Boy | 326 688 (54.8) | 235 111 (54.7) | 91 577 (54.8) |
| Girl | 269 946 (45.2) | 194 387 (45.3) | 75 559 (45.2) |
| **Language\*** |  |  |  |
| Finnish or Swedish | 546 692 (91.6) | 384 049 (89.4) | 162 643 (97.3) |
| Other | 49 942 (8.4) | 45 449 (10.6) | 4 493 (2.7) |
| **Occupational status of a parent\*\*** |  |  |  |
| Upper-level non-manual employed | 143855 (24.1) | 79 456 (18.5) | 64 399 (38.5) |
| Other forms of employment | 357 789 (60.0) | 268 802 (62.6) | 88 987 (53.2) |
| Student | 30 940 (5.2) | 26 884 (6.3) | 4 056 (2.4) |
| Unknown | 34 607 (5.8) | 28 438 (6.6) | 6 169 (3.7) |
| Unemployed | 29 443 (4.9) | 25 918 (6.0) | 3 525 (2.1) |
| **Medical history** |  |  |  |
| Recurrent acute otitis media*** | 83845 (14.1) | 46 990 (10.9) | 36 855 (22.1) |
| At least one atopic condition**** | 138 172 (23.2) | 94 267 (21.9) | 43 905 (26.3) |
| Asthma | 37 744 (6.3) | 29 405 (6.8) | 8 339 (5.0) |
| Atopic eczema | 78 776 (13.2) | 49 068 (11.4) | 29 708 (17.8) |
| Allergic rhinitis | 26203 (4.4) | 15 388 (3.6) | 10 815 (6.5) |
| Immunological deficiencies | 25 989 (4.4) | 22 147 (5.2) | 3 842 (2.3) |
| Down syndrome | 2 559 (0.4) | 2 394 (0.6) | 165 (0.1) |
| **Doctor's specialty** |  |  |  |
| No specialty | 349 280 (58.5) | 325 677 (75.8) | 23 603 (14.1) |
| Otorhinolaryngologist | 69 652 (11.7) | 7 646 (1.8) | 62 006 (37.1) |
| General practitioner | 80 185 (13.4) | 72 135 (16.8) | 8 050 (4.8) |
| Pediatrician | 81 683 (13.7) | 14 387 (3.3) | 67 296 (40.3) |
| Other | 15 834 (2.7) | 9 653 (2.2) | 6 181 (3.7) |
| **Season** |  |  |  |
| Summer | 96 085 (16.1) | 68 444 (15.9) | 27 641 (16.5) |
| Spring | 130 908 (21.9) | 104 060 (24.2) | 26 848 (16.1) |
| Fall | 188 116 (31.5) | 120 294 (28.0) | 67 822 (40.6) |
| Winter | 181 525 (30.4) | 136 700 (31.8) | 44 825 (26.8) |
| **Diagnose-group** |  |  |  |
| H65 or H67 | 77 211 (12.9) | 54 575 (12.7) | 22 636 (13.5) |
| H66 | 519 423 (87.1) | 374 923 (87.3) | 144 500 (86.5) |

\*A proxy for non-domestic-language background
\*\*According to the parent who had the higher status.
\*\*\* within a year
\*\*\*\*Allergic rhinitis (J30), atopic eczema (L20), immunological deficiencies (D80-D89), Down syndrome (Q90) or asthma (J45-J46) diagnosed prior visit.

### Management strategy

Of all the index visits (*n* = 596 634), 77.6% (n = 462 808) involved antibiotic prescription. In the crude model, treatment in the private sector was associated with lower odds of prescribing antibiotics (72.3% vs. 79.6%). However, in the adjusted model, the private sector was associated with higher odds for prescribing antibiotics (OR: 1.45; 95% CI: 1.41–1.49); due to the higher proportion of otorhinolaryngologists (who were associated with lower odds for prescribing antibiotics) in the private sector (**Figure 4**; eTable 1 in **Supplement 1**).

### Management failure

*Management failure* occurred in 7.0% of the visits and was more common in the private sector (10.8% vs. 5.2%). In most cases failure occurred in the same sector as the index visit (97.3%). The private sector had higher odds for management failure (aOR: 1.52; 95% CI: 1.48–1.56), with the physician specialty and recurrent AOM accounting for most of the attenuation (**Figure 4**; eTable 2). *Early management failure* occurred in 1.3% of the visits being more frequent in the private sector (1.9% vs. 1.1%) However, after adjustment, the sector did not have a significant effect (aOR: 1.04; 95% CI: 0.95–1.13; **Figure 4**, eTable 3).

### Antibiotic guideline adherence

Overall, 87.3% of index-visit antibiotic prescriptions were guideline adherent. Although unadjusted adherence was similar in the private and public sectors (86.3% vs 87.7%), adjusted analyses showed lower odds of guideline-adherent prescribing in the private sector (aOR, 0.64; 95% CI, 0.60–0.69; **Figure 4**; eTable 4).

### Antibiotic treatment strategy

Antibiotic selection differed by sector, with the private sector more often selecting broader-spectrum agents. Relative to amoxicillin, private-sector visits had higher odds (32.8% vs. 8.3%; aOR: 3.00; 95% credibility interval: 2.91–3.10) of amoxicillin-clavulanic acid and non-first-line antibiotics (13.7% vs. 12.3%; aOR: 1.58; 95% credibility interval: 1.52–1.65); with physician specialty (otorhinolaryngologist) accounting for most of attenuation (**Figure 4**; eTable 5).

Among non-first-line antibiotics, trimethoprim-sulfamethoxazole was most common (private 41.7% [*n* = 6873]; public 45.4%, [*n* = 19 098]). Macrolides were more common in the private sector (32.2% [*n* = 5315] vs. 23.0% [*n* = 9656]), whereas suboptimal-efficacy agents, particularly cephalexin, were more frequent in the public sector (30.5% [*n* = 12 811] vs. 25.5% [*n* = 4200]). Potentially harmful antibiotics were rarely prescribed (**Figure 3**).

**Figure 3.**
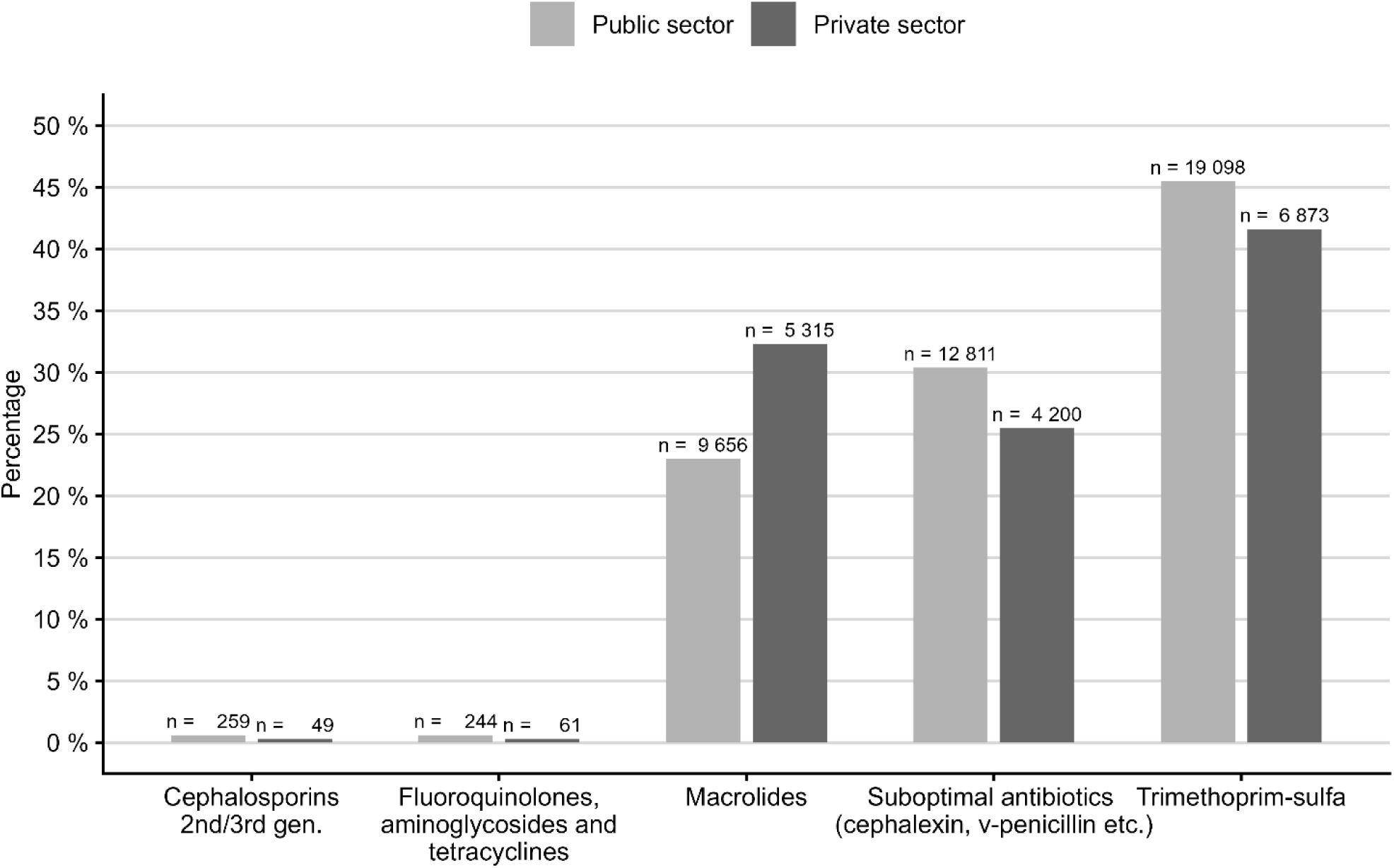
The distribution of non-first-line antibiotics by sector.

### Antibiotic treatment failure

*Antibiotic treatment failure* occurred in 7.4% of the visits with an antibiotic prescription and was more common in the private sector (11.8% vs 5.5%). The private sector had higher odds for antibiotic treatment failure (aOR, 1.48; 95% CI, 1.43–1.52; **Figure 4**; eTable 6). *Early antibiotic treatment failure* occurred in 1.3% of the visits with an antibiotic prescription and was similarly more frequent in the private sector (1.9% vs 1.1%; aOR, 1.14; 95% CI, 1.03–1.26) (**Figure 4**; eTable 7).

**Figure 4.**
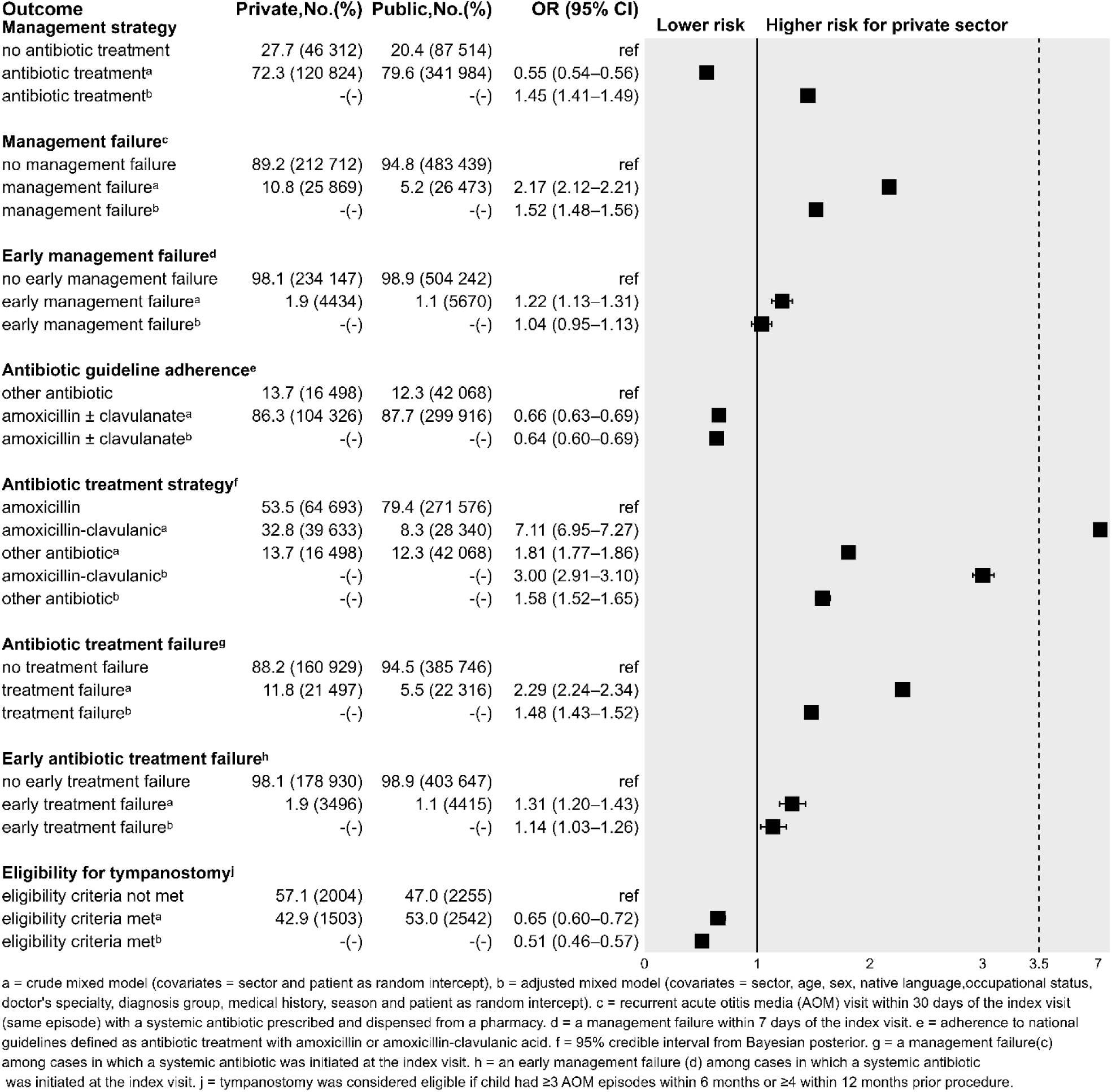
The results. OR = odds ratio; 95% CI = 2.5th–97.5th percentile confidence interval.

### Eligibility for tympanostomy-tube placement

Overall, clinical criteria for tympanostomy tube placement were met in 48.7% of procedures (53.0% in the public sector and 42.9% in the private sector). The private sector had lower odds for meeting the clinical criteria (aOR: 0.51; 95% CI: 0.46–0.57; **Figure 4**; eTable 8).

### Sensitivity analyses

Estimates for management failure, including assignment by the antibiotic-prescribing sector, and for management strategy remained largely consistent across alternative episode definitions and prescription linkage windows, although the association for management strategy was attenuated with a 14-day episode window. Estimates for tympanostomy eligibility were similar to the primary analysis (eTables 9–11 in **Supplement 1**).

## Discussion

In this nationwide registry study, we used an episode-based approach to examine public-private differences in AOM management in Finland. Adherence to recommended first-line therapy was high in both sectors, but clinically meaningful differences emerged in antibiotic selection and treatment failure rates. AOM episodes initiated in the private sector were more likely to involve antibiotic prescribing, broader-spectrum agents, and repeated antibiotic treatment.

### Antibiotic prescribing rates and management strategy

Antibiotics were prescribed in 77.6% of index visits, toward the upper end of previous Nordic and European estimates.^31–38^ Unlike visit-based studies,^31–33,38^ our 30-day episode definition (with 14-and 45-day sensitivity windows) captures the full clinical course of AOM, which likely raises prescribing rates while reducing misclassification of follow-up visits as new infections.

In unadjusted analyses, private-sector patients received antibiotics less often than public-sector patients. However, this association was reversed after adjustment, with the private sector showing higher odds of prescribing. The reversal was primarily driven by the higher proportion of otorhinolaryngologists in the private sector (42.1% of physicians vs. 2.3%), who prescribed antibiotics less frequently than other specialists, consistent with previous findings.^39,40^ This shows how profoundly a setting’s specialty mix can shape aggregate prescribing statistics.

### Antibiotic choice and guideline adherence

Overall, 87.3% of prescriptions were guideline adherent, although adherence was lower in the private sector. Private sector had substantially higher odds of amoxicillin-clavulanic acid over amoxicillin, with the odds remaining roughly three-fold after adjustment. Otorhinolaryngologist specialty emerged as the key driver of broader-spectrum antibiotic use, consistent with prior studies.^38–41^ Whether this reflects specialist judgement of risks and harms, awareness of beta-lactamase-producing organisms in complex cases, or a practice culture shaped by patient expectations is difficult to disentangle from register data. The high concentration of otorhinolaryngologists in the private sector may shape its prescribing culture, contributing to a systematic preference for broader-spectrum agents that persists even after the specialty is accounted for.^42–46^

Trimethoprim-sulfamethoxazole combinations were the most common non-first-line antibiotics in both sectors, followed by macrolides (WHO AWaRe Watch group antibiotics associated with respiratory pathogen resistance⁴), which were prescribed more frequently in the private sector. The public sector more often used agents with suboptimal efficacy against common AOM pathogens, particularly cephalexin. Potentially harmful antibiotics were prescribed rarely.

### Management and treatment failure

Management failure occurred in 7.0% of the episodes and was approximately twice as common in the private sector. Failures were concentrated in the 7–30-day window. This is clinically plausible: residual middle-ear effusion often persists for weeks after the acute infection resolves and, on re-examination, can be hard to distinguish from the active disease, particularly without tympanometry or pneumatic otoscopy. A lower threshold for re-consultation in the private sector, facilitated by private health insurance and easier access, may inflate apparent failure visits that do not represent genuine treatment failure.^47,48^ Thet broader-spectrum antibiotics coincided with higher failure rates reinforces evidence that amoxicillin-clavulanic acid offers no clinical advantage over amoxicillin as first-line AOM therapy.^43–45,49,50^

Although patient characteristics differed substantially between sectors, all analyses were adjusted for these factors. Case-mix differences therefore do not fully explain the observed differences in prescribing and treatment failure. Consistent with previous findings, lower thresholds for intervention and easier specialist access in the private sector may contribute to more intensive AOM management, including greater antibiotic use and follow-up visits.

### Tympanostomy tube eligibility

Overall, 51.3% of the tympanostomy tube procedures were performed for children who did not meet registry-based criteria for recurrent AOM, consistent with prior Finnish^51^ and international studies.^52–54^ Interpretation is limited by the lack of clinical detail in registries, including otoscopic findings and audiological assessments, which inform decisions and are required for guideline-adherent indication,^7^ as well as potential incomplete coverage of private-sector visits. These undocumented factors likely explain some apparently *non-indicated* procedures, but this finding warrants closer analysis of whether some tympanostomies depart from recommendations.

### Strengths and limitations

Strengths include nationwide, population-level coverage of all Finnish children, minimizing selection bias, including groups that are systematically underrepresented in insurance-based research settings. The Prescription Centre data captures virtually all prescriptions and dispensations in Finland, ensuring near-complete ascertainment of antibiotic use across both sectors. A clearly defined episode structure, supported by sensitivity analyses using alternative time windows with consistent results, provides a more clinically meaningful unit of analysis than visit-based approaches. Linking prescriptions to dispensation records further improved the measurement of actual antibiotic use.

Limitations include limited clinical detail as no data were available on symptom severity, tympanic membrane findings, allergies, or parental preferences. This restricts our ability to assess illness severity at presentation or determine whether differences reflect the underlying case mix. In particular, the lack of penicillin allergy information limits the interpretation of non-first-line antibiotic use. Private sector register coverage was also incomplete in the early study years, and some provider organizations may be disproportionately represented, introducing potential selection bias if the reporting is more complete among certain types of providers. Finally, the study period overlapped with the COVID-19 pandemic, which may have influenced respiratory infection epidemiology and healthcare utilization in sector-specific ways.

### Conclusions

In this nationwide, episode-based study of 295 064 Finnish children with AOM we found that, guideline adherence was high in Finland, but clinically meaningful sector differences were observed in antibiotic selection and rates of treatment failure. The private sector showed higher odds of antibiotics being prescribed, including three-fold use of amoxicillin-clavulanic acid, alongside more frequent treatment failure. These patterns likely reflect structural differences between the sectors rather than differences in clinical intent, and they support current recommendations favoring amoxicillin as the first-line treatment for uncomplicated AOM. Antibiotic prescribing rates were high in both sectors, with 89.2% of the prescriptions being dispensed, suggesting limited implementation of the watchful waiting approach and opportunities to reduce antibiotic use in AOM management. Over half of tympanostomy procedures were performed in children without documented guideline eligibility, and important quality-improvement target.

## Data Availability

The data were obtained from Finnish Institute for Health and Welfare (THL) under a research permit. Due to legal and confidentiality restrictions, the data cannot be made publicly available. Researchers may apply for access directly from the data controller in accordance with applicable regulations.

