## Supplement 1 for "Sectoral Differences in Pediatric Antibiotic Prescribing for Acute Otitis Media"

[1.	Primary healthcare system in Finland explained. (*Adapted and modified from [*Csonka *et al.,* 2025*], with permission.)*	2](#_Toc232710673)

[2.6	Doctor’s specialty. (Adapted and modified from [Csonka et al., 2025], with permission.)	4](#_Toc232710680)

### Supplementary material

#### Primary healthcare system in Finland explained. (*Adapted and modified from [*Csonka *et al.,* 2025*], with permission.)*

Finland’s primary care system is a central component of the country’s publicly funded healthcare system. It is built around municipality-run health centers that serve as the first point of contact for most medical issues. The system is designed to provide comprehensive, accessible, and equitable care to the population. Services are funded mainly through tax revenue and provided to residents at low cost.

In the Finnish primary care system, patients are treated by GPs, physicians specializing to be GPs, and doctors without any specialty. GPs provide preventive care, management of acute and chronic conditions, minor procedures, and referrals to specialized care when needed. Health centers also serve as training sites for medical students and doctors in specialist training (especially in general practice).

Acute common respiratory infections in children are primarily treated outside hospitals. Most visits are due to upper respiratory tract infections. In the public sector, infections treated outside the hospital are managed at municipal health centers by GPs, physicians specializing as GPs, and doctors without any specialty. If specialized pediatric care is needed, children are referred to central or university hospitals. After-hours care is usually centralized into regional emergency clinics, and more complex cases are referred to secondary care, typically provided by hospital districts or university hospitals.

The private healthcare sector also offers services for the pediatric population, especially in urban areas. Private clinics may provide faster and direct access also to specialists (without referral). In the private sector, in addition to GPs and non-specialists, acute infections in children are also treated by specialists, particularly ENT specialists and pediatricians. There are also major differences in the types of patients and illnesses treated by specialists in the public vs. private sectors. For example, in the public sector, ENT specialists typically do not treat common respiratory infections. Pediatricians and physicians specializing in becoming pediatricians treat some common infections in public hospital emergency departments, but in the private sector, common infections are more frequently treated by pediatric specialists compared to the public sector.

**The use of private sector services in Finland**

Most families who use private care have voluntary private insurance, which covers part or all the cost. Some employers also provide private healthcare benefits for employees’ children. Based on the FinLapset study findings from the 2023 report, approximately 60% of infants (in 2020) and 58% of four-year-old children (in 2018) in Finland had a private health insurance policy. These figures come from large national surveys with thousands of respondents and are considered reliable estimates for children in those age groups. In absolute terms, according to data from the Finnish Financial Supervisory Authority, there were 462,000 children with a private health insurance in Finland as of June 2022. This represents a significant portion of the total child population in Finland, especially considering that the total number of children under 15 was around 860,000 in that same period.

According to the FinLapset 2023 report,^1^ about 28% of children aged 0-3 years in Finland used private healthcare services (specifically private doctor visits) in 2021. For the 1-3-year-old age group, the percentage was slightly higher at 32%. These figures are based on the Social Insurance Institution of Finland’s (Kela) reimbursement data, which tracks private doctor visits that were at least partially reimbursed by the national health insurance system (excluding dental care).

Children in urban and suburban municipalities were significantly more likely to visit private doctors than those in rural areas. In densely populated municipalities, access to private clinics is easier and more convenient, which contributes to higher usage. In rural areas, private healthcare services are less available or may require long travel distances, which limits their use. In the highest usage areas, 50%–60% of children in certain age groups visited a private doctor in 2021. In the lowest usage areas, less than 10% of children visited a private practice. ^2^

#### Finnish healthcare registers

##### Finnish Care Register for Social and Health Care (Hilmo and Avohilmo)

The Care Register for Social and Health Care (Hilmo) is a nationwide data collection and reporting system maintained by the Finnish Institute for Health and Welfare (THL). It contains structured information on the provision of social and health care services across Finland. Hilmo consists of three interconnected registers:

- Healthcare Hilmo (terveys‑Hilmo): inpatient care, day surgery, and specialized outpatient care.
- Primary Care Outpatient Register (Avohilmo): primary care visits, occupational health care, and home care.
- Social Care Hilmo (sosiaali‑Hilmo): institutional social care and residential social services.

Hilmo data collection is currently expanding to cover private-sector outpatient health care as well. Hilmo contains detailed unit‑level and client‑level information that enables national monitoring, service evaluation, and research. Although the data elements vary slightly by register, the following categories are generally collected:

- Unique anonymized identifiers enabling longitudinal follow‑up
- Service‑unit information: TOPI‑coded service provider and unit, service type (e.g., inpatient ward, outpatient clinic, home‑care unit)
- Visit‑ and episode‑level information: Dates of admission, discharge, or visit
- Diagnoses and reasons for care: ICD‑10 coded diagnoses, reason(s) for visit or care episode, main, longtime and secondary diagnoses
- Procedures and interventions: surgical and non-surgical procedures, interventions, examinations, and treatments performed.^3^

##### Data Repositories of the Digital and Population Data Services Agency (DVV)

The Digital and Population Data Services Agency maintains several national data repositories and case registers that contain legally mandated personal information on individuals, families, buildings, and administrative matters. These datasets support identification services, population management, legal processes, and the provision of governmental digital services.

Population Information System contains core statutory population data for all persons residing in Finland:

- Personal identity code
- Name(s) and changes of name
- Date of birth, sex
- Nationality, native language
- Marital status and family relationships (parents, children, spouse)
- Residential address history and current domicile
- Immigration and emigration information
- Building, real estate, and dwelling data linked to individuals

And data related to other registers: Register of Guardianship Affairs, register of Prenuptial Agreements and Donations, register of the Right to Officiate Weddings, register of Cohabitation Affairs, individual Customer Data Repository (case management), mandate Register (Suomi.fi e‑Authorizations), certificate Data Repository, appointment and Service Registries.^4^

##### Prescription Centre data (Reseptikeskus)

The Medicines Data Repository includes several datasets containing comprehensive information on prescription medicines in Finland. These data originate from the national *Kanta Prescription Centre* and the *Social Insurance Institution of Finland (Kela)*.

Available from 1 May 2010 onward; fully comprehensive from 1 January 2017.

The Prescription Centre stores all electronic prescriptions and pharmacy dispensing records in Finland. Since all prescriptions are now issued electronically, this register provides a complete national record of:

- Electronic prescriptions: Prescriber details, medication name and ATC code, dosage instructions, validity period, and renewals
- Pharmacy dispensing: Dates and quantities dispensed, pharmacy identifier, remaining refill amounts, prescriber details, ATC code.^5^

##### Register of Social and Health Care Service Units (TOPI)

The Register of Social and Health Care Service Units (TOPI) is a national coding system used in Finland to uniquely identify service providers and service units within the social and health care system. Each TOPI code corresponds to a specific provider and the physical unit where a client has received care.

TOPI codes are used across national health and social care data systems, including the Care Register for Health Care (Hilmo) and other statutory registers maintained by the Finnish Institute for Health and Welfare (THL). They ensure consistent and accurate linkage of service‑unit‑level information in administrative and research datasets.

TOPI contains information on all service providers that report Hilmo data to THL—this includes institutions and units offering residential services, institutional care, and home‑care services.

Users can search service units by unit ID, name, municipality, postal code, or postal district. An up‑to‑date Excel extract of all registered units can be downloaded through the TOPI search application.^6^

##### Finnish Supervisory Agency (Valvira)

The Register of Healthcare and Social Welfare Professionals, maintained by the Finnish Supervisory Agency (formerly Valvira), comprises two linked registers: *Terhikki* for healthcare professionals and *Suosikki* for social welfare professionals. The register contains information on all regulated professional practice rights in Finland, including the validity and supervisory status of each license. An individual may hold multiple professional licenses simultaneously. The register does not capture employment information, meaning it does not record which license a person is actively practicing under or whether the person is currently working in a professional capacity. Access to the register data for research purposes requires a data permit under the Act on the Secondary Use of Health and Social Data.^7^

##### Doctor’s specialty. (Adapted and modified from [Csonka et al., 2025], with permission.)

*General practitioners (specialist in general practice)*

In Finland, to become a general practitioner (GP)—formally recognized as a Specialist in General Practice—a doctor must complete a structured five-year specialist training program following the six years long Licentiate of Medicine degree. The program begins with 9 months of full-time clinical work in primary healthcare, usually at a municipal health center. It also includes at least a year of hospital-based training in relevant specialties such as internal medicine, pediatrics, psychiatry, or geriatrics. The majority of the training takes place in primary care settings, where doctors work under the supervision of experienced GPs. The focus is on developing the skills needed to provide comprehensive, continuous care for patients of all ages and across a wide range of medical issues. Trainees must also complete theoretical coursework and pass a final written examination before graduating.^2^

**Ear, nose, and throat (ENT) specialists**

To become an ear, nose, and throat (ENT) specialist—formally titled a Specialist in Otorhinolaryngology in Finland—a doctor must complete a five- to six-year specialist training program following the Licentiate of Medicine degree. The training begins with a minimum of 9 months in primary healthcare, typically in a public health center, before continuing in hospital-based settings. The remainder of the program is conducted in central and university hospitals, where doctors train in ENT departments and rotate through subspecialty areas such as otology (ear disorders), rhinology (nasal and sinus conditions), laryngology (voice and throat disorders), and head and neck surgery. The training combines hands-on surgical experience with structured theoretical education. Trainees must complete mandatory courses and pass a national written examination to demonstrate their competence. ^2^

**Pediatricians**

To become a pediatrician in Finland, a doctor must complete a six-year specialist training program following the Licentiate of Medicine degree. The program begins with at least 9 months of training in primary healthcare, usually in child health clinics or general practice settings, where doctors gain experience in preventive care and early childhood health. The remainder of the training takes place in pediatric departments within central and university hospitals, where doctors rotate through various areas of general pediatrics and pediatric subspecialties such as neonatology, pediatric emergency care, and pediatric cardiology. In addition to clinical training, the program includes theoretical coursework and requires candidates to pass a national written examination. ^2^

**Physicians with no medical specialty**

In Finland, doctors who have completed the basic medical degree (Licentiate of Medicine) but have not specialized are fully licensed to practice medicine. Often referred to as non-specialist doctors or general physicians, they are authorized by Valvira, the national licensing authority, to work independently in a variety of clinical settings. These doctors typically work in health centers, hospitals, emergency departments, or occupational health services. Many are early in their careers and may work under the supervision of specialists or in multidisciplinary teams. Some take temporary or locum positions while gaining experience or considering future specialization. Although they are not specialists in a specific field, they are trained to manage common medical issues and refer patients to specialized care when needed. While many eventually enter specialist training, others continue working as general physicians, especially in roles where broad clinical skills are in demand.^2^

#### Methods

##### Study design and population

This nationwide register-based cohort study used linked individual-level data from primary (AvoHilmo^3^), secondary care (Hilmo^3^) and population registers^4,5,7,8^ All registers cover both public and private health care providers; private sector data have been available since 2015, with complete coverage from 2026.^9^

##### Participants

We included all children under 18 years of age who had at least one health care contact registered in AvoHilmo or Hilmo between January 1, 2017, to December 31, 2022, and received a diagnosis (primary, secondary or long-term) of AOM (diagnosis codes H65-H67) defined by the International Classification of Diseases, 10th Revision (ICD-10). For the sub cohort of children who underwent tympanostomy tube placement we included all children from the main cohort who underwent tympanostomy tube insertion (procedure codes SPAT1019, DCA20) between January 1, 2020, to December 31 2022 defined by Nordic Classification of Surgical Procedures (NCSP).

##### ICD-10 codes

*Non-suppurative otitis media*: H65.0, H65.1, H65.2, H65.3, H65.4, H65.9

*Suppurative otitis media*: H66.0, H66.1, H66.2, H66.3, H66.4, H66.9

*Otitis media linked to other diseases*: H67.0, H67.1, H67.8

##### Data processing

We first identified AOM visits from Hilmo and AvoHilmo between 01-01-2017 and 31-12-2022 using ICD-10 codes H65- H67. Sociodemographic data from Digital and Population Data Services Agency and Statistics Finland were linked, and the cohort was restricted to individuals under 18 years of age. ^3,4,8^ The responsible physician was identified using pseudonymized physician registration number. Physician specialty from Finnish Supervisory Agency was linked using the identifiers and the corresponding calendar year to ensure that specialty information reflected the physician’s status at the time of the visit.^7^ Finally, antibiotic prescriptions (ATC code: J01*) and dispensations from the national Prescription Centre^5^were linked with the AOM visits using a pseudonymized patient identifier and the visit date. Antibiotic prescriptions were linked to the AOM visit if issued within two days following the visit (Fig. 2b). As a sensitivity analysis, we also evaluated zero and seven-day windows to link the visit with a prescription.

AOM visits were grouped into AOM episodes. An episode index visit was defined as either the patient's first recorded AOM visit or any subsequent AOM visit occurring ≥ 30 after the index visit, thereby marking the beginning of a new AOM episode. AOM visits occurring within 30 days of an index visit were considered being a part of the same episode and classified as repetitive visits. The 30-day window is clinically justified, as middle ear effusion typically resolves within approximately one month. ^10^

To evaluate eligibility to tympanostomy, we identified a sub cohort of patients who had undergone tympanostomy between 01-01-2020 and 31-12-2022 using NCSP procedure codes DCA20 and SPAT1019. This study period was used to ensure better coverage of private sector visits. Then patient and physician-level information were linked to tympanostomy procedures as described above.

Each visit generates as many rows as there are prescriptions associated with that visit. For each physician, all antibiotic prescriptions were retained, and the least favorable antibiotic choice per physician was used as the outcome for analysis (in order: non-first-line, amoxicillin-clavulanate, amoxicillin). A pipeline of data processing is presented in efigure 1 below.

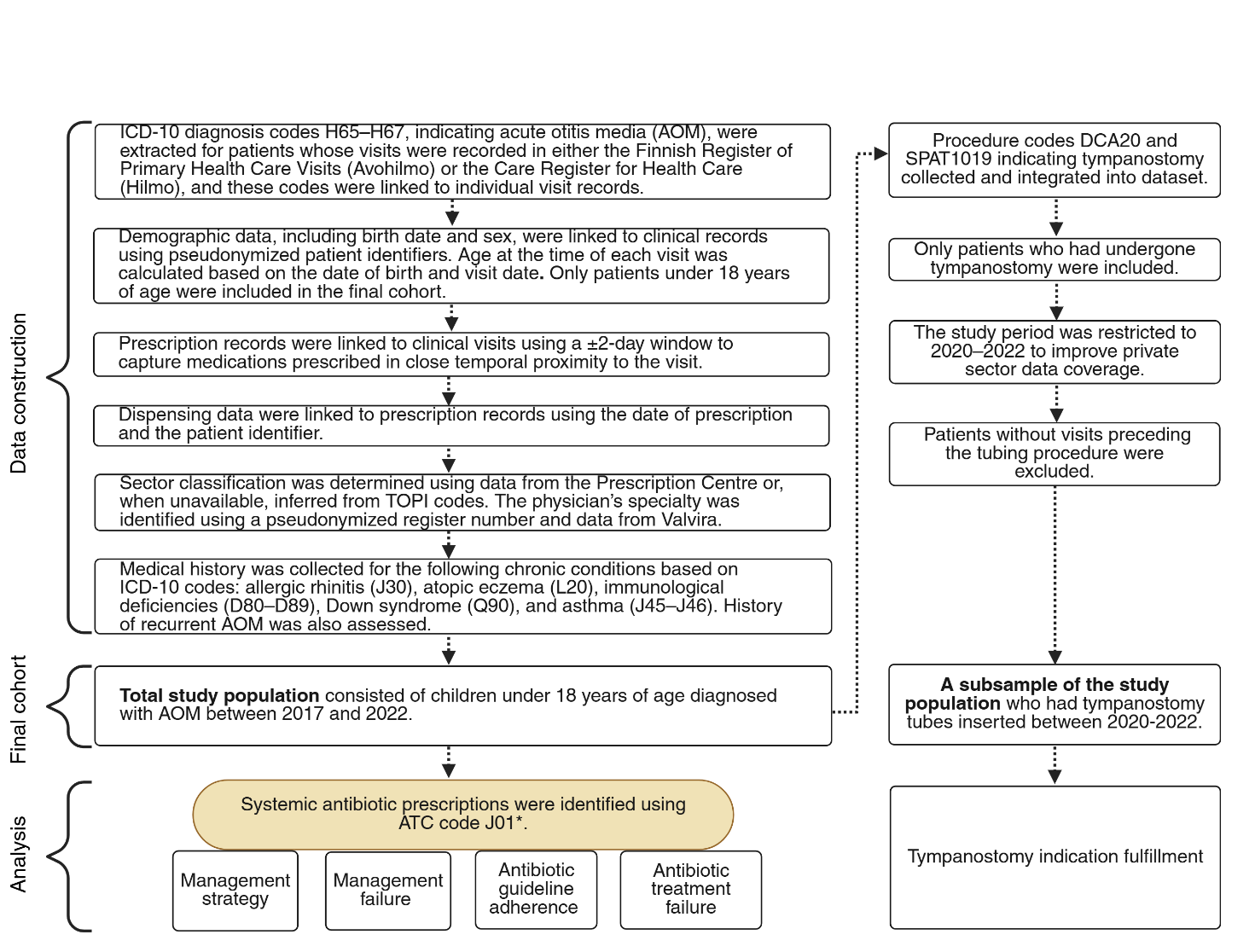

***eFigure 1*** ***(biorender) Data processing pipeline*** *(Created in https://BioRender.com).*

AOM=Acute otitis media, ATC = Anatomical Therapeutic Chemical code, TOPI codes=location register codes

##### Covariates

The patient’s birth date was used to calculate the age at the time of the visit, and patients were classified into 4 groups: <2 years, 2-4.9 years, 5-11.9 years and 12-17.9 years. In the sub-cohort analysis, the two oldest age groups were combined due to the small number of patients in the 12–17.9-year age group (n = 306).

Sex was coded as girl or boy. Parental occupational status was based on the higher-status parent and categorized as upper-level non-manual, other forms of employment (including retirees, n = 5162), unemployed, student, or unknown. Native language was categorized as Finnish, Swedish, or other (proxy for non-domestic-language background).

Patient’s medical history was derived from diagnosis records by ICD-10 codes and linked to AOM visit data using the pseudonymized personal identity code. Only diagnoses recorded prior to or on the same day as the AOM visit were included. Conditions included were allergic rhinitis (J30), atopic eczema (L20), immunological deficiencies (D80–D89), Down syndrome (Q90), and asthma (J45–J46). ^11–13^

Recurrent otitis media was defined as ≥3 episodes within a six-month period or ≥4 within one year, and treated as a background factor for one year after the last qualifying episode. Healthcare sector was categorized as *public* or *private* by provider type, determined from prescription data or, when unavailable and for the tympanostomy subgroup, from location register (TOPI) codes.^6^ Doctor’s specialty, per the Finnish Supervisory Agency^7^, was classified as *otorhinolaryngologist, general practicer, pediatrician, other specialty,* or *no specialty*.

We included diagnosis category as a covariate, grouping diagnoses into H66 (suppurative or unspecified otitis media) and H65/H67 (non-suppurative otitis media and otitis associated with other diseases) based on clinical relevance and sample size considerations. Season was assigned from the month of the visit: *winter* (December–February), *spring* (March–May), *summer* (June–August), and *fall* (September–November).^14^

##### ATC-codes of non-first-line antibiotics

*Second-line antibiotics:*

Macrolides: J01FA*,

Trimethoprim-sulfa: J01EE*

Cephalosporines 2^nd^. gen.: J01DC*

Cephalosporines third gen.: J01DD*

*Sub-optimal antibiotics:*

Cephalosporines 1^st^ gen. J01DB*

V-pen, G-pen: J01CE*

Clindamycin: J01FF01

Pivmecillinam: J01CA08

Flucloxacillin: J01CF05

Trimethoprim: J01EA*

Nitrofurantoin: J01XE01

*Potentially harmful (due the known adverse-effects):*

Tetracyclines: J01AA*

Aminoglycosides: J01GB*

Fluoroquinolones: J01M*

##### Statistical analysis

###### **General linear mixed-effects model (GLMM)**

We evaluated the association between healthcare sector and study outcomes using both standard logistic regression models (GLM) and generalized linear mixed-effects models (GLMM), with patient identifier (FID) included as a random intercept to account for repeated observations within individuals.^15,16^

We compared models with Akaike Information Criterion (AIC) and GLMMs consistently provided better or comparable fit relative to GLMs. Although the GLM yielded a slightly lower AIC for eligibility for tympanostomy, effect estimates were virtually identical to those from the corresponding GLMM (differences <0.1), and mixed-effects models were therefore retained for consistency across analyses.

###### **Bayesian multinominal model (BMM)**

We fitted both the crude and adjusted Bayesian multinomial models using Stan via the brms interface with CmdStanR backend. We run each model with 4 chains of 700 warmup and 700 sampling iterations (1400 total per chain), using within-chain parallelization across 3 threads. We defined priors empirically based on a frequentist multinomial logistic regression model (*nnet::multinom*) fitted to the same dataset, using amoxicillin as the reference category. This approach anchors priors to observed data patterns while allowing the posterior to update freely.

We specified priors for regression coefficients in three tiers based on effect magnitude and consistency. Strongly informative priors (normal (μ, 1.0)) were assigned to covariates with large and consistent effects, including *private sector care, otorhinolaryngology specialty*, and history of *recurrent AOM*. Prior means (μ) were set to 1.0, 1.6, and 1.0, respectively, reflecting the magnitude of the corresponding frequentist estimates. We used moderately informative priors (normal (μ, 0.7)) for covariates with smaller or less consistent effects, including *pediatric specialty, older age group* (12–18 years), *other specialties, atopic conditions*, and *diagnosis group*.

For *diagnosis group* (H66), an asymmetric prior mean of −0.4 was specified to reflect the consistently negative association observed in the frequentist model. For all remaining covariates, including *sex, native language, socioeconomic status, and season*, we assigned weakly informative priors centered at zero (normal (0, 0.5)), reflecting small or inconsistent effects. We set intercept priors to normal (-1.2, 1.5) for *amoxicillin-clavulanic* and normal (-1.8, 1.5) for *other antibiotics*, based on frequentist intercept estimates of -3.17 and -1.82, with sufficiently wide variance to allow the data to dominate. Priors for the standard deviation of the random intercept (patient-level effects) were left at their default weakly informative specification in brms (Student-t distribution with 3 degrees of freedom, mean 0, and scale 2.5), providing regularization while allowing sufficient flexibility.

We assessed convergence using the potential scale reduction factor (R̂), effective sample size (ESS), and visual inspection of trace plots. In the adjusted model, all fixed-effect parameters had R̂ = 1.00. The random-effect standard deviations had R̂ values of 1.01 (amoxicillin-clavulanic) and 1.01 (other antibiotics), both within the accepted threshold (R̂ < 1.05). Bulk effective sample sizes were 509 and 684 for the two random-effect standard deviations, respectively and tail ESS values were 1205 and 1129. Trace plots indicated adequate mixing across chains, and no divergent transitions were observed after warmup.

#### Sensitivity analyses

###### **Management failure by sector that prescribed antibiotics at recurrent visit**

In the primary analysis, management failures were assigned to the sector of the index visit. As a sensitivity analysis, we reassigned failures to the sector prescribing antibiotics at the failure visit, regardless of the index-visit sector, to assess whether attribution of failures influenced the results.

###### **Episode Interval**

###### The primary analysis defined otitis media episodes using a 30-day interval. To assess the robustness of this definition, two alternative episode intervals were evaluated: a 14-day interval and a 45-day interval. We re-estimated management failure and management strategy outcomes under each definition using the same mixed-model specifications as in the primary analysis. Results were consistent across all three episode interval definitions. The odds ratio for management failure in the private sector was 2.06 (95% CI, 2.01–2.12) with the 14-day interval and 1.98 (95% CI, 1.95–2.02) with the 45-day interval, compared with 2.17 (95% CI, 2.12–2.21) in the primary analysis. For management strategy, the odds ratio for antibiotic treatment in the private sector was 0.48 (95% CI, 0.47–0.49) with the 14-day interval and 0.61 (95% CI, 0.60–0.62) with the 45-day interval. Because the 14-day interval produced the largest deviation from the primary analysis, an adjusted model was additionally fitted. The adjusted estimate (aOR, 1.25; 95% CI, 1.21–1.28) was directionally consistent with the primary adjusted analysis, although the magnitude of the association was somewhat attenuated (primary analysis: aOR, 1.45; 95% CI, 1.41–1.49).

###### **Prescription Linkage Window**

The primary analysis linked index visits to antibiotic prescriptions with 2-day linkage window. To evaluate whether a broader linkage window affected results, we applied a 7-day linkage window, allowing prescriptions dispensed within 7 days of the index visit to be attributed to that visit. Under the 7-day window, the odds ratio for management failure in the private sector was 2.15 (95% CI 2.11–2.20) and for antibiotic treatment strategy 0.57 (95% CI 0.56–0.58), both consistent with the primary analysis estimates.

###### **Tympanostomy Eligibility with 14-days episode intervall**

We assessed eligibility for tympanostomy additionally using a 14-day episode interval in place of the primary 30-day interval, to examine whether episode definition affected the classification of recurrent acute otitis media and subsequent tympanostomy eligibility. Results remained consistent with the primary analysis (aOR 0.59 95% CI 0.53–0.66), althought rates of meeting the criteria were larger in both sectors (57.8% in private sector and 63.5 in public sector).

Overall, findings were robust across all sensitivity analyses, supporting the stability of the primary estimates with respect to episode interval definition, prescription linkage window, and tympanostomy eligibility criteria. See results in eTables 8.1-8.3.

#### Results of multivariable generalized linear mixed models

##### eTable 1 Management strategy

|  | **Crude mixed model** | | | **Adjusted mixed model** | | |
| --- | --- | --- | --- | --- | --- | --- |
|  | Odds ratio | 2.5 % | 97.5 % | Odds ratio | 2.5 % | 97.5 % |
| Public sector | ref |  |  | ref |  |  |
| Private sector | 0.55 | 0.54 | 0.56 | 1.45 | 1.41 | 1.49 |
| Age group: 0-2 years |  |  |  | ref |  |  |
| Age group: 2-5 years |  |  |  | 1.02 | 1.00 | 1.04 |
| Age group: 5-12 years |  |  |  | 1.11 | 1.08 | 1.13 |
| Age group: 12-18 years |  |  |  | 1.23 | 1.18 | 1.28 |
| Sex: boy |  |  |  | ref |  |  |
| Sex: girl |  |  |  | 1.06 | 1.05 | 1.08 |
| Language: Finnish or Swedish |  |  |  | ref |  |  |
| Language: other |  |  |  | 1.16 | 1.12 | 1.20 |
| Occupational status*: student |  |  |  | ref |  |  |
| Occupational status*: other forms of employment |  |  |  | 0.97 | 0.93 | 1.01 |
| Occupational status*: unknown |  |  |  | 0.94 | 0.89 | 0.99 |
|  | **Crude mixed model** | | | **Adjusted mixed model** | | |
|  | Odds ratio | 2.5 % | 97.5 % | Odds ratio | 2.5 % | 97.5 % |
| Occupational status*: unemployed |  |  |  | 0.97 | 0.92 | 1.03 |
| Occupational status*: upper non-manual worker |  |  |  | 0.85 | 0.81 | 0.89 |
| Doctor's specialty: no specialty |  |  |  | ref |  |  |
| Doctor's specialty: otorhinolaryngologist |  |  |  | 0.20 | 0.19 | 0.20 |
| Doctor's specialty: general practicer |  |  |  | 2.20 | 2.13 | 2.26 |
| Doctor's specialty: other specialty |  |  |  | 1.59 | 1.50 | 1.68 |
| Doctor's specialty: pediatrician |  |  |  | 1.10 | 1.06 | 1.14 |
| Season: summer |  |  |  | ref |  |  |
| Season: spring |  |  |  | 1.17 | 1.14 | 1.20 |
| Season: autumn |  |  |  | 1.58 | 1.54 | 1.62 |
| Season: winter |  |  |  | 1.25 | 1.22 | 1.28 |
| No recurrent Acute Otitis Media within a year |  |  |  | ref |  |  |
| Recurrent Acute Otitis Media within a year |  |  |  | 0.19 | 0.19 | 0.20 |
| No atopic condition** prior visit |  |  |  | ref |  |  |
| Atopic condition** prior visit |  |  |  | 0.87 | 0.85 | 0.89 |
| Diagnose-group: H65 or H67 |  |  |  | ref |  |  |
| Diagnose-group: H66 |  |  |  | 12.01 | 11.73 | 12.31 |
| *According to the parent who had the higher status |  |  |  |  |  |  |
| **Allergic rhinitis (J30), atopic eczema (L20), immunological deficiencies (D80–D89), Down syndrome (Q90), | | | | | | |
| and asthma (J45–J46). |  |  |  |  |  |  |

##### eTable 2 Management failure

|  | **Crude mixed effects model** | | | **Adjusted mixed effects model** | | |
| --- | --- | --- | --- | --- | --- | --- |
|  | Odds Ratio | 2.5 % | 97.5 % | Odds Ratio | 2.5 % | 97.5 % |
| Public sector | ref |  |  | ref |  |  |
| Private sector | 2.17 | 2.12 | 2.21 | 1.52 | 1.48 | 1.56 |
| Age group: 0-2 years |  |  |  | ref |  |  |
| Age group: 2-5 years |  |  |  | 0.70 | 0.68 | 0.71 |
| Age group: 5-12 years |  |  |  | 0.43 | 0.42 | 0.44 |
| Age group: 12-18 years |  |  |  | 0.36 | 0.34 | 0.38 |
| Sex: boy |  |  |  | ref |  |  |
| Sex: girl |  |  |  | 0.97 | 0.95 | 0.99 |
| Language: Finnish or Swedish |  |  |  | ref |  |  |
| Language: other |  |  |  | 0.90 | 0.87 | 0.94 |
| Occupational status*: student |  |  |  | ref |  |  |
| Occupational status*: other forms of employment |  |  |  | 0.96 | 0.91 | 1.01 |
| Occupational status*: unknown |  |  |  | 0.94 | 0.89 | 1.00 |
| Occupational status*: unemployed |  |  |  | 0.95 | 0.89 | 1.01 |
| Occupational status*: upper non-manual worker |  |  |  | 0.94 | 0.89 | 0.99 |
| Doctor's specialty: no specialty |  |  |  | ref |  |  |
|  | **Crude mixed effects model** | | | **Adjusted mixed effects model** | | |
|  | Odds Ratio | 2.5 % | 97.5 % | Odds Ratio | 2.5 % | 97.5 % |
| Doctor's specialty: otorhinolaryngologist |  |  |  | 1.12 | 1.08 | 1.16 |
| Doctor's specialty: general practicer |  |  |  | 1.20 | 1.16 | 1.23 |
| Doctor's specialty: other specialty |  |  |  | 1.36 | 1.29 | 1.44 |
| Doctor's specialty: pediatrician |  |  |  | 1.36 | 1.32 | 1.41 |
| Season: summer |  |  |  | ref |  |  |
| Season: spring |  |  |  | 1.15 | 1.11 | 1.19 |
| Season: autumn |  |  |  | 1.64 | 1.59 | 1.69 |
| Season: winter |  |  |  | 1.39 | 1.35 | 1.44 |
| No recurrent Acute Otitis Media within a year |  |  |  | ref |  |  |
| Recurrent Acute Otitis Media within a year |  |  |  | 2.15 | 2.11 | 2.20 |
| No atopic condition** prior visit |  |  |  | ref |  |  |
| Atopic condition** prior visit |  |  |  | 1.07 | 1.04 | 1.09 |
| Diagnose-group: H65 or H67 |  |  |  | ref |  |  |
| Diagnose-group: H66 |  |  |  | 2.94 | 2.83 | 3.06 |
| *According to the parent who had the higher status. | |  |  |  |  |  |
| **Allergic rhinitis (J30), atopic eczema (L20), immunological deficiencies (D80–D89), Down syndrome (Q90), | | | | | | |
| and asthma (J45–J46). |  |  |  |  |  |  |

##### eTable 3 Early management failure

|  | **Crude mixed effects model** | | | **Adjusted mixed effects model** | | |
| --- | --- | --- | --- | --- | --- | --- |
|  | Odds ratio | 2.5 % | 97.5 % | Odds ratio | 2.5 % | 97.5 % |
| Public sector | ref |  |  | ref |  |  |
| Private sector | 1.22 | 1.13 | 1.31 | 1.04 | 0.95 | 1.13 |
| Age group: 0-2 years |  |  |  | ref |  |  |
| Age group: 2-5 years |  |  |  | 0.78 | 0.73 | 0.85 |
| Age group: 5-12 years |  |  |  | 0.70 | 0.63 | 0.78 |
| Age group: 12-18 years |  |  |  | 0.76 | 0.63 | 0.92 |
| Sex: boy |  |  |  | ref |  |  |
| Sex: girl |  |  |  | 0.97 | 0.89 | 1.07 |
| Language: Finnish or Swedish |  |  |  | ref |  |  |
| Language: other |  |  |  | 0.88 | 0.74 | 1.05 |
| Occupational status*: student |  |  |  | ref |  |  |
| Occupational status*: other forms of employment |  |  |  | 0.91 | 0.76 | 1.09 |
| Occupational status*: unknown |  |  |  | 0.97 | 0.78 | 1.22 |
| Occupational status*: unemployed |  |  |  | 0.87 | 0.69 | 1.09 |
| Occupational status*: upper non-manual worker |  |  |  | 1.01 | 0.83 | 1.22 |
| Doctor's specialty: no specialty |  |  |  | ref |  |  |
| Doctor's specialty: otorhinolaryngologist |  |  |  | 1.14 | 1.04 | 1.25 |
| Doctor's specialty: general practicer |  |  |  | 1.05 | 0.96 | 1.15 |
| Doctor's specialty: other specialty |  |  |  | 1.48 | 1.27 | 1.72 |
| Doctor's specialty: pediatrician |  |  |  | 1.34 | 1.22 | 1.46 |
| Season: summer |  |  |  | ref |  |  |
|  | **Crude mixed effects model** | | | **Adjusted mixed effects model** | | |
|  | Odds ratio | 2.5 % | 97.5 % | Odds ratio | 2.5 % | 97.5 % |
| Season: spring |  |  |  | 1.04 | 0.94 | 1.14 |
| Season: autumn |  |  |  | 1.30 | 1.20 | 1.42 |
| Season: winter |  |  |  | 1.20 | 1.10 | 1.31 |
| No recurrent Acute Otitis Media within a year |  |  |  | ref |  |  |
| Recurrent Acute Otitis Media within a year |  |  |  | 1.30 | 1.22 | 1.39 |
| No atopic condition** prior visit |  |  |  | ref |  |  |
| Atopic condition** prior visit |  |  |  | 1.13 | 1.03 | 1.24 |
| Diagnose-group: H65 or H67 |  |  |  | ref |  |  |
| Diagnose-group: H66 |  |  |  | 4.07 | 3.64 | 4.56 |
| *According to the parent who had the higher status. | |  |  |  |  |  |
| **Allergic rhinitis (J30), atopic eczema (L20), immunological deficiencies (D80–D89), Down syndrome (Q90), | | | | | | |
| and asthma (J45–J46). |  |  |  |  |  |  |

##### eTable 4 Guideline adherence

|  | **Crude mixed effects model** | | | **Adjusted mixed effects model** | | |
| --- | --- | --- | --- | --- | --- | --- |
|  | Odds ratio | 2.5 % | 97.5 % | Odds ratio | 2.5 % | 97.5 % |
| Public sector |  |  |  | ref |  |  |
| Private sector | 0.66 | 0.63 | 0.69 | 0.64 | 0.60 | 0.69 |
| Age group: 0-2 years |  |  |  | ref |  |  |
| Age group: 2-5 years |  |  |  | 0.93 | 0.89 | 0.98 |
| Age group: 5-12 years |  |  |  | 0.96 | 0.91 | 1.02 |
| Age group: 12-18 years |  |  |  | 0.56 | 0.51 | 0.61 |
| Sex: boy |  |  |  | ref |  |  |
| Sex: girl |  |  |  | 1.02 | 0.97 | 1.08 |
| Language: Finnish or Swedish |  |  |  | ref |  |  |
| Language: other |  |  |  | 1.45 | 1.29 | 1.63 |
| Occupational status*: student |  |  |  | ref |  |  |
| Occupational status*: other forms of employment |  |  |  | 0.65 | 0.60 | 0.71 |
| Occupational status*: unknown |  |  |  | 0.70 | 0.67 | 0.74 |
| Occupational status*: unemployed |  |  |  | 0.58 | 0.53 | 0.64 |
| Occupational status*: upper non-manual worker |  |  |  | 1.52 | 1.41 | 1.63 |
| Doctor's specialty: no specialty |  |  |  | ref |  |  |
| Doctor's specialty: otorhinolaryngologist |  |  |  | 1.01 | 0.92 | 1.12 |
| Doctor's specialty: general practicer |  |  |  | 1.03 | 0.91 | 1.18 |
| Doctor's specialty: other specialty |  |  |  | 1.08 | 0.95 | 1.23 |
| Doctor's specialty: pediatrician |  |  |  | 1.26 | 1.12 | 1.40 |
| Season: summer |  |  |  | ref |  |  |
| Season: spring |  |  |  | 0.87 | 0.82 | 0.92 |
| Season: autumn |  |  |  | 1.10 | 1.04 | 1.16 |
| Season: winter |  |  |  | 0.85 | 0.80 | 0.90 |
| No recurrent Acute Otitis Media within a year |  |  |  | ref |  |  |
|  | **Crude mixed effects model** | | | **Adjusted mixed effects model** | | |
|  | Odds ratio | 2.5 % | 97.5% | Odds ratio | 2.5 % | 97.5% |
| Recurrent Acute Otitis Media within a year |  |  |  | 0.30 | 0.28 | 0.32 |
| No atopic condition** prior visit |  |  |  | ref |  |  |
| Atopic condition** prior visit |  |  |  | 0.91 | 0.86 | 0.96 |
| Diagnose-group: H65 or H67 |  |  |  | ref |  |  |
| Diagnose-group: H66 |  |  |  | 2.66 | 2.48 | 2.85 |
| *According to the parent who had the higher status |  |  |  |  |  |  |
| **Allergic rhinitis (J30), atopic eczema (L20), immunological deficiencies (D80–D89), Down syndrome (Q90), | | | | | |  |
| and asthma (J45–J46). |  |  |  |  |  |  |

##### eTable 5 Antibiotic treatment strategy: Bayesian multinominal model

|  | **Crude model** | | | **Adjusted model** | | |
| --- | --- | --- | --- | --- | --- | --- |
| **Amoxicillin-clavulanic** | OR | 2.5 % | 97.5 % | OR | 2.5 % | 97.5 % |
| Public sector | ref |  |  | ref |  |  |
| Private sector | 7.11 | 6.95 | 7.27 | 3.00 | 2.91 | 3.10 |
| Age group: 0-2 years |  |  |  | ref |  |  |
| Age group: 2-5 years |  |  |  | 1.19 | 1.16 | 1.22 |
| Age group: 5-12 years |  |  |  | 1.09 | 1.06 | 1.12 |
| Age group: 12-18 years |  |  |  | 1.50 | 1.43 | 1.57 |
| Sex: boy |  |  |  | ref |  |  |
| Sex: girl |  |  |  | 0.92 | 0.90 | 0.94 |
| Language: Finnish or Swedish |  |  |  | ref |  |  |
| Language: other |  |  |  | 0.83 | 0.80 | 0.87 |
| Occupational status*: student |  |  |  | ref |  |  |
| Occupational status*: other forms of employment |  |  |  | 0.97 | 0.92 | 1.02 |
| Occupational status*: unknown |  |  |  | 0.88 | 0.82 | 0.94 |
| Occupational status*: unemployed |  |  |  | 0.89 | 0.83 | 0.95 |
| Occupational status*: upper non-manual worker |  |  |  | 0.92 | 0.88 | 0.98 |
| Doctor's specialty: no specialty |  |  |  | ref |  |  |
| Doctor's specialty: otorhinolaryngologist |  |  |  | 5.99 | 5.74 | 6.24 |
| Doctor's specialty: general practicer |  |  |  | 1.15 | 1.12 | 1.19 |
| Doctor's specialty: other specialty |  |  |  | 1.43 | 1.35 | 1.51 |
| Doctor's specialty: pediatrician |  |  |  | 2.25 | 2.18 | 2.33 |
| Diagnose-group: H65 or H67 |  |  |  | ref |  |  |
| Diagnose-group: H66 |  |  |  | 0.81 | 0.78 | 0.85 |
| No recurrent Acute Otitis Media within a year |  |  |  | ref |  |  |
| Recurrent Acute Otitis Media within a year |  |  |  | 2.88 | 2.78 | 2.98 |
| Comorbidity** prior visit |  |  |  | ref |  |  |
| Comorbidity** prior visit |  |  |  | 1.08 | 1.06 | 1.11 |
| Season: summer |  |  |  | ref |  |  |
| Season: spring |  |  |  | 0.95 | 0.92 | 0.99 |
| Season: autumn |  |  |  | 0.96 | 0.93 | 0.99 |
| Season: winter |  |  |  | 1.00 | 0.97 | 1.03 |
| **Other** | **OR** | **2.5 %** | **97.5 %** | **OR** | **2.5 %** | **97.5 %** |
|  | **Crude model** | | | **Adjusted model** | | |
| **Other** | **OR** | **2.5 %** | **97.5 %** | **OR** | **2.5 %** | **97.5 %** |
| Public sector | ref |  |  | ref |  |  |
| Private sector | 1.81 | 1.77 | 1.86 | 1.58 | 1.52 | 1.65 |
| Age group: 0-2 years |  |  |  | ref |  |  |
| Age group: 2-5 years |  |  |  | 1.08 | 1.05 | 1.11 |
| Age group: 5-12 years |  |  |  | 1.21 | 1.17 | 1.25 |
| Age group: 12-18 years |  |  |  | 1.86 | 1.77 | 1.96 |
| Sex: boy |  |  |  | ref |  |  |
| Sex: girl |  |  |  | 0.95 | 0.92 | 0.97 |
| Language: Finnish or Swedish |  |  |  | ref |  |  |
| Language: other |  |  |  | 0.63 | 0.59 | 0.66 |
| Occupational status*: student |  |  |  | ref |  |  |
| Occupational status*: other forms of employment |  |  |  | 1.01 | 0.96 | 1.07 |
| Occupational status*: unknown |  |  |  | 0.94 | 0.87 | 1.01 |
| Occupational status*: unemployed |  |  |  | 0.97 | 0.90 | 1.04 |
| Occupational status*: upper non-manual worker |  |  |  | 0.81 | 0.76 | 0.86 |
| Doctor's specialty: no specialty |  |  |  | ref |  |  |
| Doctor's specialty: otorhinolaryngologist |  |  |  | 2.47 | 2.33 | 2.61 |
| Doctor's specialty: general practicer |  |  |  | 1.24 | 1.21 | 1.28 |
| Doctor's specialty: other specialty |  |  |  | 1.66 | 1.56 | 1.76 |
| Doctor's specialty: pediatrician |  |  |  | 0.78 | 0.75 | 0.82 |
| Diagnose-group: H65 or H67 |  |  |  | ref |  |  |
| Diagnose-group: H66 |  |  |  | 0.47 | 0.45 | 0.49 |
| No recurrent Acute Otitis Media within a year |  |  |  | ref |  |  |
| Recurrent Acute Otitis Media within a year |  |  |  | 4.11 | 3.95 | 4.28 |
| Comorbidity** prior visit |  |  |  | ref |  |  |
| Comorbidity** prior visit |  |  |  | 1.18 | 1.15 | 1.22 |
| Season: summer |  |  |  | ref |  |  |
| Season: spring |  |  |  | 1.07 | 1.03 | 1.11 |
| Season: autumn |  |  |  | 0.88 | 0.85 | 0.91 |
| Season: winter |  |  |  | 1.02 | 0.98 | 1.05 |
| **Amoxicillin** | ref | ref | ref | ref | ref | ref |
| *According to the parent who had the higher status. |  |  |  |  |  |  |
| **Allergic rhinitis (J30***), atopic eczema (L20***), immunological deficiencies (D80–D89***), | | | | | | |
| Down syndrome (Q90***), and asthma (J45–J46***). | |  |  |  |  |  |
| OR = crude odds ratio; with Bayesian 2.5th–97.5th percentile credible intervals | | | | | | |

##### eTable 6 Antibiotic treatment failure

|  | **Crude mixed effects model** | | | **Adjusted mixed effects model** | | |
| --- | --- | --- | --- | --- | --- | --- |
|  | Odds ratio | 2.5 % | 97.5 % | Odds ratio | 2.5 % | 97.5 % |
| Public sector | ref |  |  | ref |  |  |
| Private sector | 2.29 | 2.24 | 2.34 | 1.48 | 1.43 | 1.52 |
| Age group: 0-2 years |  |  |  | ref |  |  |
| Age group: 2-5 years |  |  |  | 0.70 | 0.68 | 0.71 |
| Age group: 5-12 years |  |  |  | 0.43 | 0.41 | 0.44 |
| Age group: 12-18 years |  |  |  | 0.36 | 0.33 | 0.38 |
| Sex: boy |  |  |  | ref |  |  |
| Sex: girl |  |  |  | 0.97 | 0.95 | 0.99 |
|  | **Crude mixed effects model** | | | **Adjusted mixed effects model** | | |
|  | Odds ratio | 2.5 % | 97.5 % | Odds ratio | 2.5 % | 97.5 % |
| Language: Finnish or Swedish |  |  |  | ref |  |  |
| Language: other |  |  |  | 0.87 | 0.83 | 0.90 |
| Occupational status*: student |  |  |  | ref |  |  |
| Occupational status*: other forms of employment |  |  |  | 0.97 | 0.92 | 1.02 |
| Occupational status*: unknown |  |  |  | 0.95 | 0.89 | 1.02 |
| Occupational status*: unemployed |  |  |  | 0.94 | 0.87 | 1.01 |
| Occupational status*: upper non-manual worker |  |  |  | 0.96 | 0.91 | 1.02 |
| Doctor's specialty: no specialty |  |  |  | ref |  |  |
| Doctor's specialty: otorhinolaryngologist |  |  |  | 1.39 | 1.33 | 1.44 |
| Doctor's specialty: general practicer |  |  |  | 1.07 | 1.04 | 1.11 |
| Doctor's specialty: other specialty |  |  |  | 1.22 | 1.15 | 1.30 |
| Doctor's specialty: pediatrician |  |  |  | 1.28 | 1.24 | 1.33 |
| Season: summer |  |  |  | ref |  |  |
| Season: spring |  |  |  | 1.10 | 1.06 | 1.14 |
| Season: autumn |  |  |  | 1.51 | 1.46 | 1.57 |
| Season: winter |  |  |  | 1.31 | 1.27 | 1.36 |
| No recurrent Acute Otitis Media within a year |  |  |  | ref |  |  |
| Recurrent Acute Otitis Media within a year |  |  |  | 2.25 | 2.19 | 2.30 |
| No atopic condition** prior visit |  |  |  | ref |  |  |
| Atopic condition** prior visit |  |  |  | 1.08 | 1.05 | 1.11 |
| Diagnose-group: H65 or H67 |  |  |  | ref |  |  |
| Diagnose-group: H66 |  |  |  | 1.57 | 1.50 | 1.64 |
| *According to the parent who had the higher status. | |  |  |  |  |  |
| **Allergic rhinitis (J30), atopic eczema (L20), immunological deficiencies (D80–D89), Down syndrome (Q90), | | | | | | |
| and asthma (J45–J46). |  |  |  |  |  |  |

##### eTable 7 Early antibiotic treatment failure

|  | **Crude mixed effects model** | | | **Adjusted mixed effects model** | | |
| --- | --- | --- | --- | --- | --- | --- |
|  | Odds ratio | 2.5 % | 97.5 % | Odds ratio | 2.5 % | 97.5 % |
| Public sector | ref |  |  | ref |  |  |
| Private sector | 1.31 | 1.20 | 1.43 | 1.14 | 1.03 | 1.26 |
| Age group: 0-2 years |  |  |  | ref |  |  |
| Age group: 2-5 years |  |  |  | 0.81 | 0.74 | 0.89 |
| Age group: 5-12 years |  |  |  | 0.74 | 0.66 | 0.84 |
| Age group: 12-18 years |  |  |  | 0.76 | 0.61 | 0.96 |
| Sex: boy |  |  |  | ref |  |  |
| Sex: girl |  |  |  | 0.98 | 0.88 | 1.09 |
| Language: Finnish or Swedish |  |  |  | ref |  |  |
| Language: other |  |  |  | 0.87 | 0.70 | 1.07 |
| Occupational status*: student |  |  |  | ref |  |  |
| Occupational status*: other forms of employment |  |  |  | 0.92 | 0.74 | 1.14 |
| Occupational status*: unknown |  |  |  | 0.98 | 0.75 | 1.28 |
| Occupational status*: unemployed |  |  |  | 0.88 | 0.66 | 1.16 |
| Occupational status*: upper non-manual worker |  |  |  | 1.05 | 0.84 | 1.33 |
| Doctor's specialty: no specialty |  |  |  | ref |  |  |
|  | **Crude mixed effects model** | | | **Adjusted mixed effects model** | | |
|  | Odds ratio | 2.5 % | 97.5 % | Odds ratio | 2.5 % | 97.5 % |
| Doctor's specialty: otorhinolaryngologist |  |  |  | 1.22 | 1.09 | 1.37 |
| Doctor's specialty: general practicer |  |  |  | 0.88 | 0.79 | 0.97 |
| Doctor's specialty: other specialty |  |  |  | 1.25 | 1.05 | 1.49 |
| Doctor's specialty: pediatrician |  |  |  | 1.12 | 1.01 | 1.24 |
| Season: summer |  |  |  | ref |  |  |
| Season: spring |  |  |  | 0.96 | 0.86 | 1.08 |
| Season: autumn |  |  |  | 1.17 | 1.06 | 1.30 |
| Season: winter |  |  |  | 1.11 | 1.00 | 1.24 |
| No recurrent Acute Otitis Media within a year |  |  |  | ref |  |  |
| Recurrent Acute Otitis Media within a year |  |  |  | 1.19 | 1.10 | 1.29 |
| No atopic condition** prior visit |  |  |  |  |  |  |
| Atopic condition** prior visit |  |  |  | 1.16 | 1.04 | 1.30 |
| Diagnose-group: H65 or H67 |  |  |  | ref |  |  |
| Diagnose-group: H66 |  |  |  | 2.24 | 1.94 | 2.59 |
| *According to the parent who had the higher status. | |  |  |  |  |  |
| **Allergic rhinitis (J30), atopic eczema (L20), immunological deficiencies (D80–D89), Down syndrome (Q90), | | | | | | |
| and asthma (J45–J46). |  |  |  |  |  |  |

##### eTable 8 Eligibility for tympanostomy

|  | **Crude mixed effects model** | | | **Adjusted mixed effects model** | | |
| --- | --- | --- | --- | --- | --- | --- |
|  | Odds ratio | 2.5 % | 97.5 % | Odds ratio | 2.5 % | 97.5 % |
| Public sector | ref |  |  | ref |  |  |
| Private sector | 0.65 | 0.60 | 0.72 | 0.51 | 0.46 | 0.57 |
| Age group: 0-2 years |  |  |  | ref |  |  |
| Age group: 2-5 years |  |  |  | 0.89 | 0.80 | 1.00 |
| Age group: 5-18 years |  |  |  | 0.32 | 0.28 | 0.37 |
| Sex: boy |  |  |  | ref |  |  |
| Sex: girl |  |  |  | 1.00 | 0.91 | 1.10 |
| Language: Finnish or Swedish |  |  |  | ref |  |  |
| Language: other |  |  |  | 0.76 | 0.61 | 0.95 |
| Socioeconomic status*: student |  |  |  | ref |  |  |
| Socioeconomic status*: other forms of employment | |  |  | 1.17 | 0.90 | 1.51 |
| Socioeconomic status*: unknown |  |  |  | 1.12 | 0.81 | 1.55 |
| Socioeconomic status*: unemployed |  |  |  | 1.33 | 0.96 | 1.84 |
| Socioeconomic status*: upper non-manual worker |  |  |  | 1.19 | 0.91 | 1.56 |
| Season: summer |  |  |  | ref |  |  |
| Season: spring |  |  |  | 0.76 | 0.64 | 0.89 |
| Season: autumn |  |  |  | 0.94 | 0.80 | 1.10 |
| Season: winter |  |  |  | 0.69 | 0.59 | 0.8 |
| No comorbidity** prior visit |  |  |  | ref |  |  |
| Comorbidity** prior visit |  |  |  | 1.67 | 1.49 | 1.87 |
| *According to the parent who had the higher status |  |  |  |  |  |  |
| **Allergic rhinitis (J30), atopic eczema (L20), immunological deficiencies (D80–D89), Down syndrome (Q90), | | | | | |  |
| and asthma (J45–J46). |  |  |  |  |  |  |

#### Sensitivity analyses

##### eTable 9 Management Failure and Management Strategy

| **A. Prescription Linkage Window** | | | | | | | |
| --- | --- | --- | --- | --- | --- | --- | --- |
|  | **0-day linkage window** | | |  | **7-day linkage window** | | |
|  | **OR** | **2.5 %** | **97.5 %** |  | **OR** | **2.5 %** | **97.5 %** |
| ***Management Failure*** | | | | | | | |
| Private Index sector | 2,249 | 2,206 | 2,293 |  | 2,154 | 2,113 | 2,195 |
| ***Management Strategy*** | | | | | | | |
| Private Sector | 0,620 | 0,608 | 0,633 |  | 0,565 | 0,555 | 0,575 |
| **B. Episode Interval** | | | | | | | |
|  | **14-day episode interval** | | |  | **45-day episode interval** | | |
|  | **OR** | **2.5 %** | **97.5 %** |  | **OR** | **2.5 %** | **97.5 %** |
| ***Management Failure*** | | | | | | | |
| Private Index Sector | 2,063 | 2,009 | 2,119 |  | 1,982 | 1,947 | 2,017 |
| ***Management Strategy*** | | | | | | | |
| Private Sector | 0,477* | 0,469 | 0,485 |  | 0,610 | 0,597 | 0,623 |
| *OR = crude odds ratio; 95% CI = 2.5th–97.5th percentile confidence interval. aOR= adjusted odds ratio*  **aOR= 1.25(1.21-1.28)* | | | | | | | |

##### eTable 10 Management failure by the sector that prescribed antibiotic at the time of the failure

|  | **Crude mixed effects model** | | | **Adjusted mixed effects model** | | |
| --- | --- | --- | --- | --- | --- | --- |
|  | Odds ratio | 2.5% | 97.5% | Odds ratio | 2.5% | 97.5% |
| Public sector | ref |  |  | ref |  |  |
| Private sector | 2.32 | 2.28 | 2.37 | 1.82 | 1.76 | 1.87 |
| Age group: 0-2 years |  |  |  | ref |  |  |
| Age group: 2-5 years |  |  |  | 0.69 | 0.68 | 0.71 |
| Age group: 5-12 years |  |  |  | 0.43 | 0.42 | 0.44 |
| Age group: 12-18 years |  |  |  | 0.36 | 0.34 | 0.38 |
| Sex: boy |  |  |  | ref |  |  |
| Sex: girl |  |  |  | 0.97 | 0.95 | 0.99 |
| Language: Finnish or Swedish |  |  |  | ref |  |  |
| Language: other |  |  |  | 0.92 | 0.88 | 0.95 |
| Socioeconomic status*: student |  |  |  | ref |  |  |
| Socioeconomic status*: other forms of employment |  |  |  | 0.95 | 0.91 | 1.00 |
| Socioeconomic status*: unknown |  |  |  | 0.94 | 0.89 | 1.00 |
| Socioeconomic status*: unemployed |  |  |  | 0.95 | 0.890 | 1.02 |
| Socioeconomic status*: upper non-manual worker |  |  |  | 0.93 | 0.88 | 0.98 |
| Doctor's specialty: no specialty |  |  |  | ref |  |  |
|  | **Crude mixed effects model** | | | **Adjusted mixed effects model** | | |
|  | Odds ratio | 2.5% | 97.5% | Odds ratio | 2.5% | 97.5% |
| Doctor's specialty: otorhinolaryngologist |  |  |  | 0.94 | 0.91 | 0.98 |
| Doctor's specialty: general practicer |  |  |  | 1.19 | 1.15 | 1.22 |
| Doctor's specialty: other specialty |  |  |  | 1.26 | 1.19 | 1.33 |
| Doctor's specialty: pediatrician |  |  |  | 1.18 | 1.14 | 1.22 |
| Season: summer |  |  |  | ref |  |  |
| Season: spring |  |  |  | 1.16 | 1.12 | 1.20 |
| Season: autumn |  |  |  | 1.63 | 1.58 | 1.68 |
| Season: winter |  |  |  | 1.40 | 1.36 | 1.45 |
| No recurrent Acute Otitis Media within a year |  |  |  | ref |  |  |
| Recurrent Acute Otitis Media within a year |  |  |  | 2.16 | 2.11 | 2.20 |
| No atopic condition** prior visit |  |  |  | ref |  |  |
| Atopic condition** prior visit |  |  |  | 1.06 | 1.04 | 1.09 |
| Diagnose-group: H65 or H67 |  |  |  | ref |  |  |
| Diagnose-group: H66 |  |  |  | 2.93 | 2.82 | 3.05 |
| *According to the parent who had the higher status | | | | | | |
| **Allergic rhinitis (J30), atopic eczema (L20), immunological deficiencies (D80–D89), Down syndrome (Q90), | | | | | | |
| and asthma (J45–J46). | | | | | | |

##### eTable 11 Eligibility for tympanostomy when recurrent acute otitis media is calculated with 14-day episode interval.

|  | **Crude mixed effects model** | | | **Adjusted mixed effects model** | | |
| --- | --- | --- | --- | --- | --- | --- |
|  | Odds ratio | 2.5 % | 97.5 % | Odds ratio | 2.5 % | 97.5 % |
| Public sector | ref |  |  | ref |  |  |
| Private sector | 0.78 | 0.71 | 0.86 | 0.59 | 0.53 | 0.66 |
| Age group: 0-2 years |  |  |  | ref |  |  |
| Age group: 2-5 years |  |  |  | 0.77 | 0.69 | 0.86 |
| Age group: 5-18 years |  |  |  | 0.27 | 0.23 | 0.31 |
| Sex: boy |  |  |  | ref |  |  |
| Sex: girl |  |  |  | 1.00 | 0.91 | 1.11 |
| Language: Finnish or Swedish |  |  |  | ref |  |  |
| Language: other |  |  |  | 0.78 | 0.62 | 0.98 |
| Occupational status*: student |  |  |  | ref |  |  |
| Occupational status*: other forms of employment |  |  |  | 1.15 | 0.89 | 1.50 |
| Occupational status*: unknown |  |  |  | 1.05 | 0.75 | 1.47 |
| Occupational status*: unemployed |  |  |  | 1.19 | 0.85 | 1.67 |
| Occupational status*: upper non-manual worker |  |  |  | 1.13 | 0.86 | 1.48 |
| No comorbidity** prior visit |  |  |  | ref |  |  |
| Comorbidity** prior visit |  |  |  | 1.69 | 1.50 | 1.90 |
| Season: summer |  |  |  | ref |  |  |
| Season: spring |  |  |  | 0.64 | 0.54 | 0.76 |
| Season: autumn |  |  |  | 1.02 | 0.86 | 1.20 |
| Season: winter |  |  |  | 0.68 | 0.58 | 0.79 |
| *According to the parent who had the higher status | | | | | | |
| **Allergic rhinitis (J30), atopic eczema (L20), immunological deficiencies (D80–D89), Down syndrome (Q90), | | | | | | |
| and asthma (J45–J46). | | | | | | |
